# High prescribing and state-level variation in z-drug use among Medicare patients

**DOI:** 10.1101/2022.05.10.22274909

**Authors:** Kaitlin E. Anderson, James L. Basting, Rachel I. Gifeisman, Donovan J. Harris, Antonica R. Rajan, Kenneth L. McCall, Brain J. Piper

## Abstract

**Background:** Z-drugs are nonbenzodiazepine hypnotics used for sleep initiation and maintenance that have been shown to increase the risk of fall-related injuries in patients aged 65 and older. The American Geriatrics Society Beers criteria classifies them as a high-risk medication and strongly recommends avoiding prescribing z-drugs to the elderly due to adverse effects. The objective of this study was to determine the prevalence of Z-drug prescribing among Medicare patients.

**Methods:** Z-drug prescription data was extracted from the Centers for Medicare and Medicaid Services State Drug Utilization Data (CMS SDUD) for 2018. For all 50 states, the number of prescriptions per 100 Medicare enrollees and days-supply per prescription was determined. The percentage of total prescriptions prescribed by each specialty and the average number of prescriptions prescribed by providers within each specialty was also determined.

**Results:** Zolpidem was the most prescribed z-drug, making up 95.0% of all z-drug prescriptions. Prescriptions per 100 enrollees were significantly elevated in Utah (28.2) and Arkansas (26.7) and significantly lower in Hawaii (9.3) relative to the national average (17.5). The specialties family medicine (32.1%), internal medicine (31.4%), and psychiatry (11.7%) made up the largest percentages of total prescriptions. The number of prescriptions per provider was significantly elevated for psychiatry relative to other specialties.

**Conclusions:** Contrary to the Beers criteria, z-drugs are being prescribed to Medicare enrollees over age 65 at high rates. While sleep disturbances in the elderly should not be ignored, alternative therapies must be considered to avoid the serious adverse effects of z-drugs.

**Key Points:** More than one-half million Medicare patients received z-drug prescriptions in 2018 that were inconsistent with the Beers criteria.

Z-drug prescriptions per 100,000 Medicare patients were significantly elevated in Utah and Arkansas.

Family Medicine had the highest number of prescriptions out of all medical specialties.

Psychiatry had a significantly higher number of prescriptions per provider compared to all other specialties.

## Introduction

Over one-third (35%) of the US population experiences short sleep, less than 7 hours [1]. Up to 50% of people over 60 years report sleep disorder symptoms while 12-20% of the elderly were diagnosed with insomnia disorder [1]. Symptoms include difficulty initiating and maintaining sleep, fatigue, mood disturbances and impaired daytime performance [2]. The elderly are considered a “special population” as they are at an increased risk for adverse drug effects and comorbidities including heart disease, stroke, diabetes, depression, and cancer [3].

Nonbenzodiazepine hypnotics zolpidem, zaleplon, zopiclone, and eszopiclone, commonly referred to as ‘z-drugs’ represent a large class of sedatives which are approved for sleep initiation and maintenance. While most z-drugs act as a selective agonist at the GABA_A_ α1 subunit, zopiclone and eszopiclone are non-selective and bind to the α_1_, α_2_, α_3_ and α_5_ subunits (i.e., the same mechanism as benzodiazepines) [4]. Z-drugs have been shown to increase the risk of fall-related injuries, such as fractures and traumatic brain injuries, in elderly patients [5]. Additionally, withdrawal symptoms for these Schedule IV drugs include delirium, which can potentiate the risk of fall-related injuries. These adverse effects are reflected in the American Geriatrics Society (AGS) Beers criteria guidelines for these drugs, which made a strong recommendation in 2019 to avoid prescribing z-drugs to patients 65 years and older.

Interestingly, the AGS also reports that z-drugs provide only minimal improvement in sleep duration and latency in the elderly [6]. Similarly, Kaiser-Permanente considers z-drugs as not safer for older adults than benzodiazepines and are a “high-risk medication in the elderly” [7].

Therefore, it is important to track the prescription patterns of these drugs to patients aged 65 or older in the United States. This study provides a nationwide examination of z-drug prescriptions to Medicare enrollees in 2018.

## Methods

### Procedures

Medicare Part D data was acquired from the Centers for Medicare and Medicaid Services State Drug Utilization Data (CMS SDUD) for 2018 [8]. Both generic and name brand formulations of z-drugs were considered for analysis (zolpidem, Ambien, Edluar, Intermezzo, Zolpimist, eszopiclone, Lunesta, zopiclone, zaleplon, and Sonata, Supplemental Table 1).

### Data-analysis

For each state, z-drug prescriptions were summed and divided by the number of Medicare enrollees in that state, as reported by the CMS. This data was normalized and reported as the number of Z-drug prescriptions per 100 Medicare enrollees. States that were > +1.96 standard deviations outside the mean were categorized as statistically significant (*p* < .05). An exploratory correlation was completed between percent obesity in 2018 [9] and prescriptions per state. Additionally, the total days’ supply was divided by the total number of prescriptions for each state and reported as days-supply per prescription. An analysis of providers prescribing z-drugs was also performed. Prescriber data was obtained from the Medicare Part D Prescriber Dataset from CMS [8]. Using a python script (Supplemental Appendix 2), the sum of the total claim counts for each specialty was calculated and used to determine the percentage of all z-drug prescriptions prescribed by each specialty. Additionally, the total number of z-drug prescriptions within each specialty with greater than 100 providers was divided by the number of providers within that specialty as reported in CMS. Specialties with a prescriptions-per-provider value >1.96 standard deviations outside the mean were identified as statistically significant (*p* < .05).

## Results

Overall, zolpidem accounted for the vast preponderance (95.0%) of z-drug prescriptions. Additionally, generic formulations made up 99.7% of prescriptions.

The average number prescriptions per 100 Medicare enrollees was 17.5 + 4.0. There was a three-fold (3.04) difference between the highest and lowest states. Utah (28.2) and Arkansas (26.7) were significantly elevated, and Hawai’i (9.3) was significantly lower relative to the state mean (Figure 1). States with higher levels of obesity also had significantly more prescriptions (*r*(49) = +0.35, *p* < .05, Figure 2).

**Figure 1.**
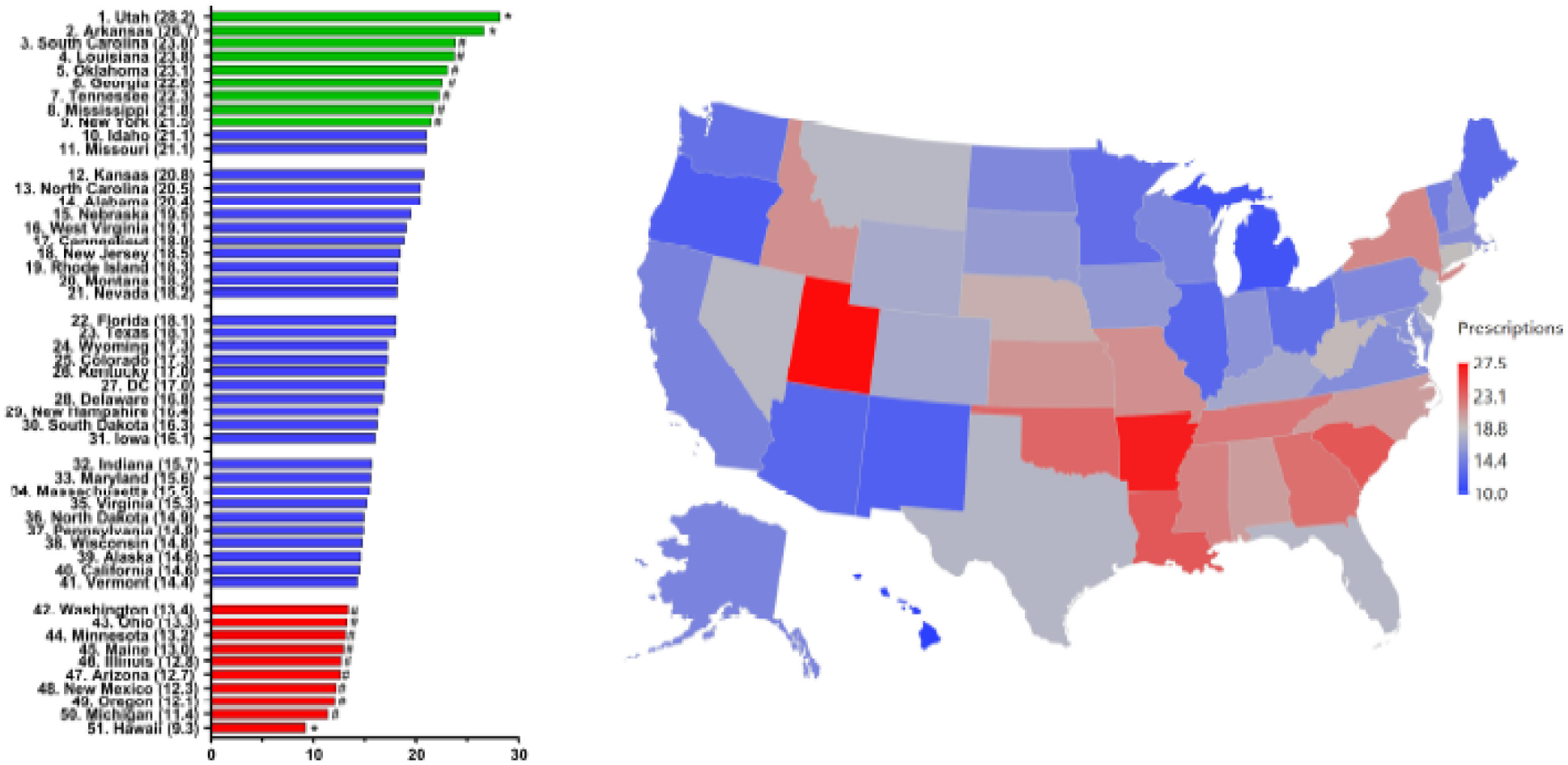
Z-drug prescriptions per 100 Medicare enrollees ranked by state. States with a * are > 1.96 SD and # were > 1.50 SD.

**Figure 2.**
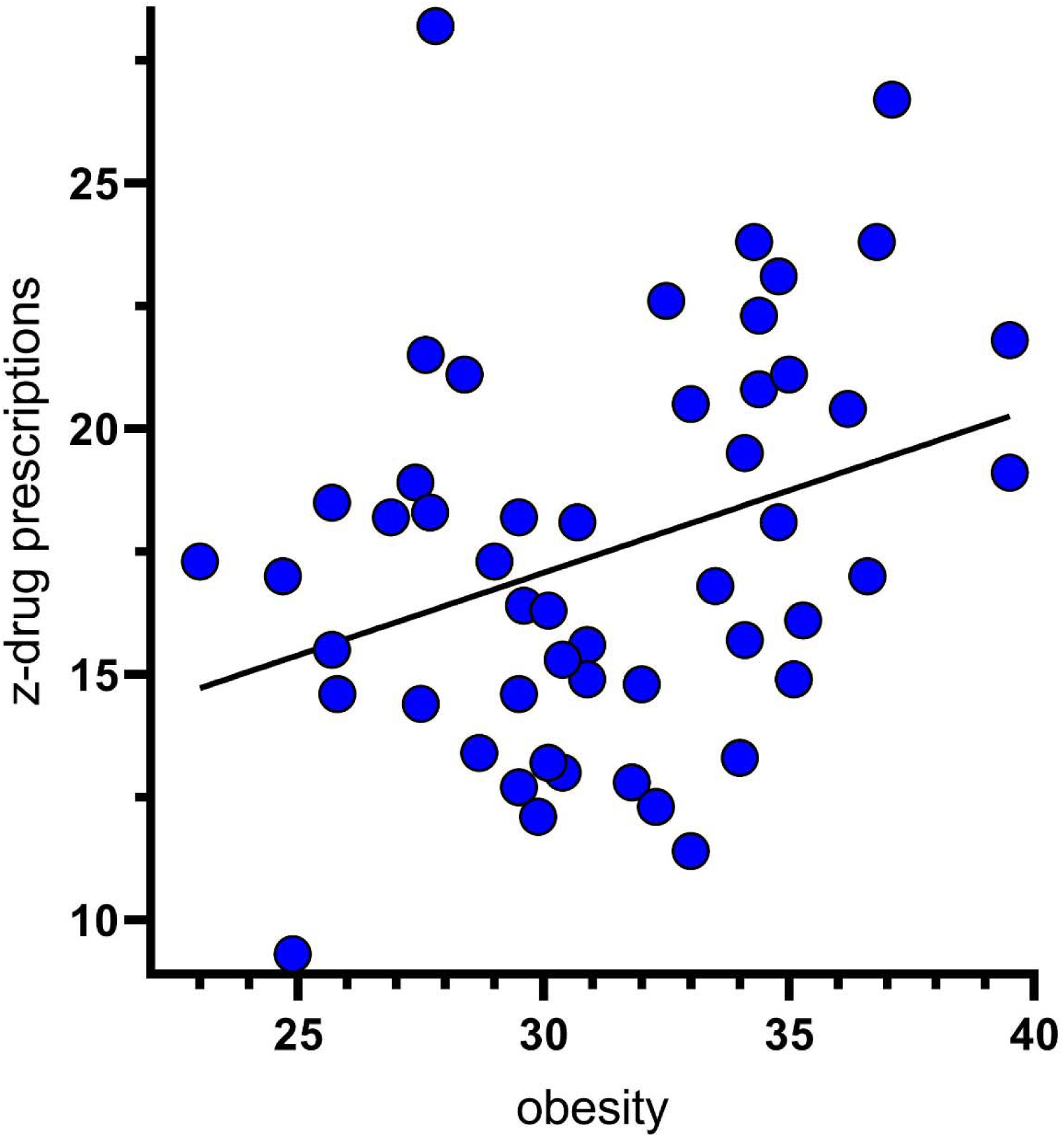
Scatterplot showing significant association between percent obesity and Z-drug prescriptions Medicare patients per state (R^2^(49) = .106, *p* = .02)

The average days-supply per prescription was 34.9. Delaware (40.2) was significantly longer, and New York (29.3) was significantly shorter than the national mean (Supplemental Figure 1). States with more prescriptions per enrollee tended to have shorter days’ supply but this was not significant (*r*(51) = 0.26, *p* = .063).

Three medical specialties comprised three-quarters (75.2%) of all z-drug prescriptions in 2018. Family Medicine (2,351,301, 32.1%) had the highest number of prescriptions followed by internal medicine (2,298,487, 31.4%) and psychiatry (853,649, 11.7%). All other specialties combined comprised 24.8% (Supplemental Figure 2). The average number of z-drug prescriptions per provider was 40.3 + 12.1. Psychiatry was significantly elevated relative to the average with 76.5 prescriptions per provider (Figure 3).

**Figure 3.**
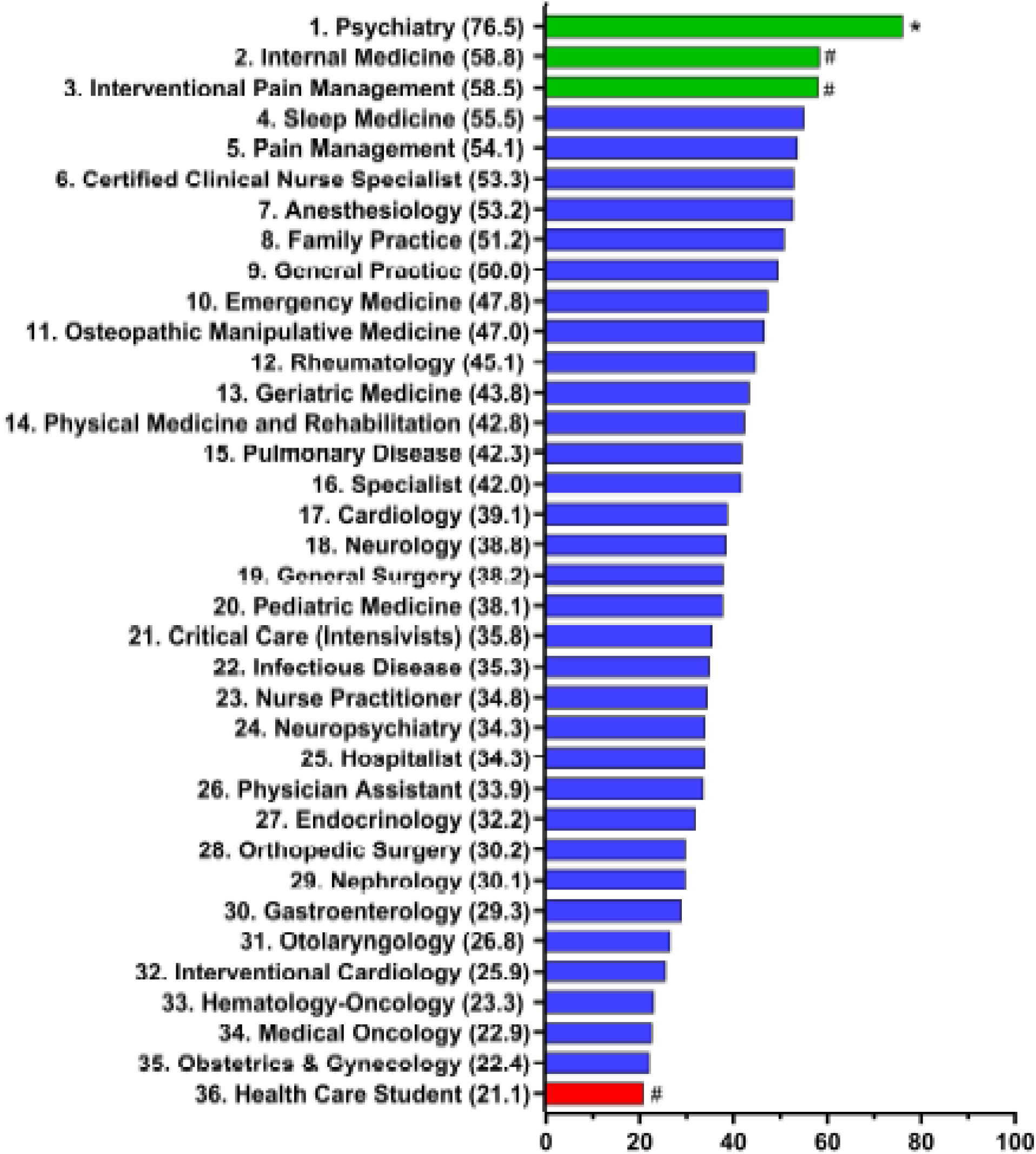
Z-drug prescriptions by specialty to Medicare patients in 2018. * are > 1.96 SD and # were > 1.50 SD.

## Discussion

It is clear Medicare patients are being prescribed z-drugs, predominantly zolpidem, at high rates despite clear guidelines to avoid use of these drugs in the elderly by the AGS due to the increased risk of adverse effects including fractures from falls, stroke, and psychological distress [6]. While elderly patients are being prescribed these drugs nationwide, there also appears to be substantial state level differences in z-drug prescriptions. We are skeptical that the identified three-fold difference between states in prescribing rates is matched by a three-fold difference in the prevalence of sleep problems. We also found a low, but significant, correlation (r = .33) between the prevalence of obesity and the z-drug prescribing rate. Obesity is a contributor to sleep apnea, and this can result in complaints to prescribers of unsatisfactory sleep quality. A systematic review identified some evidence that eszopiclone could improve continuous positive airway pressure adherence [10]. Utah and Arkansas had significantly elevated z-drug prescriptions per 100 Medicare enrollees. Meanwhile, Hawaii had the lowest number of z-drug prescriptions per 100 Medicare enrollees. This is consistent with other data showing that Hawaii also has the lowest rates of opioid and antibiotic prescriptions [11, 12]. Potential explanations for this include that Hawaii is among the healthiest states when ranked by rates of obesity, insurance coverage, and numbers of preventable hospitalizations [13]. Additionally, the large Asian population is more likely to be skeptical of prescription drugs – opting instead for increased familial support and alternative medicine [14]. However, more research is necessary to understand which factors are most influential in keeping Hawaii’s z-drug prescriptions low, as well as why Arkansas and Utah’s prescriptions are so comparatively high.

In the analysis of z-drug prescriptions per specialty; family medicine, internal medicine, and psychiatry took the largest share of prescriptions. The predominance of family medicine and internal medicine over psychiatry may be explained by patients with sleep problems presenting more often to their primary care physician rather than a specialist provider [15]. This disparity may also be influenced by only about 23% of US psychiatrists being covered by Medicare [16]. However, it did not escape notice that one hundred percent of the working group for the American Psychiatric Association’s Diagnostic and Statistical Manual for Sleep/Wake Disorders had ties to the pharmaceutical industry which raises concerns about further relaxation of the diagnostic criteria in future revisions of the DSM-5-TR [17]. Other studies have documented general practitioners (GPs) mixed perspectives on z-drug prescription, noting that many GPs experienced tension between their desire to help patients with insomnia and their fear of contributing to over-prescription of drugs with such potentially severe adverse effects [18].

It is important to emphasize that, as of 2018, less than 10% of Medicare patients were less than age 65 [19]. In addition, less than 3% of Medicare enrollees received hospice care in 2018 [20]. Based on the volume and duration of prescriptions, more than one-half million Medicare enrollees received z-drug prescriptions in 2018 that were inconsistent with the Beers Criteria. Further research with other databases including electronic health records will be necessary to identify additional subsets of Medicare patients where guidelines were not applicable, as well as determination of patient subgroups (e.g., nursing home residents and those who are obese) at greatest risk of receiving potentially inappropriate z-drug prescriptions.

## Conclusion

Sleep disorders in the elderly remain a significant problem and should not be ignored. Considering that guidelines in the United States state z-drug use is contraindicated in the elder population due to the risk of adverse events, especially falls, it is important to begin implementing changes in prescription practices such as utilizing alternative therapies. Some treatment options that have proven successful include cognitive behavior therapy or the development of consistent, healthy sleeping habits. Perhaps more drastic measures may be appropriate if patients remain symptomatic, but alternative therapies should certainly be attempted initially. Further research is needed to ascertain exactly whether these alternative therapies are not being initiated and why regional differences exist in number of prescriptions of z-drugs per individual prescribers.

## Supporting information

Zdrug_providers

## Data Availability

All data produced in the present work are contained in the manuscript as associated supplemental materials.

https://www.cms.gov/Research-Statistics-Data-and-Systems/Statistics-Trends-and-Reports/Medicare-Provider-Charge-Data/Part-D-Prescriber

## Acknowledgements

BJP was supported by the Health Resources Services Administration (D34HP31025). Software used in this effort was provided by the NIEHS (T32-ES007060-31A1). Iris Johnston provided technical support.

---

**Supplemental Figure 1.**
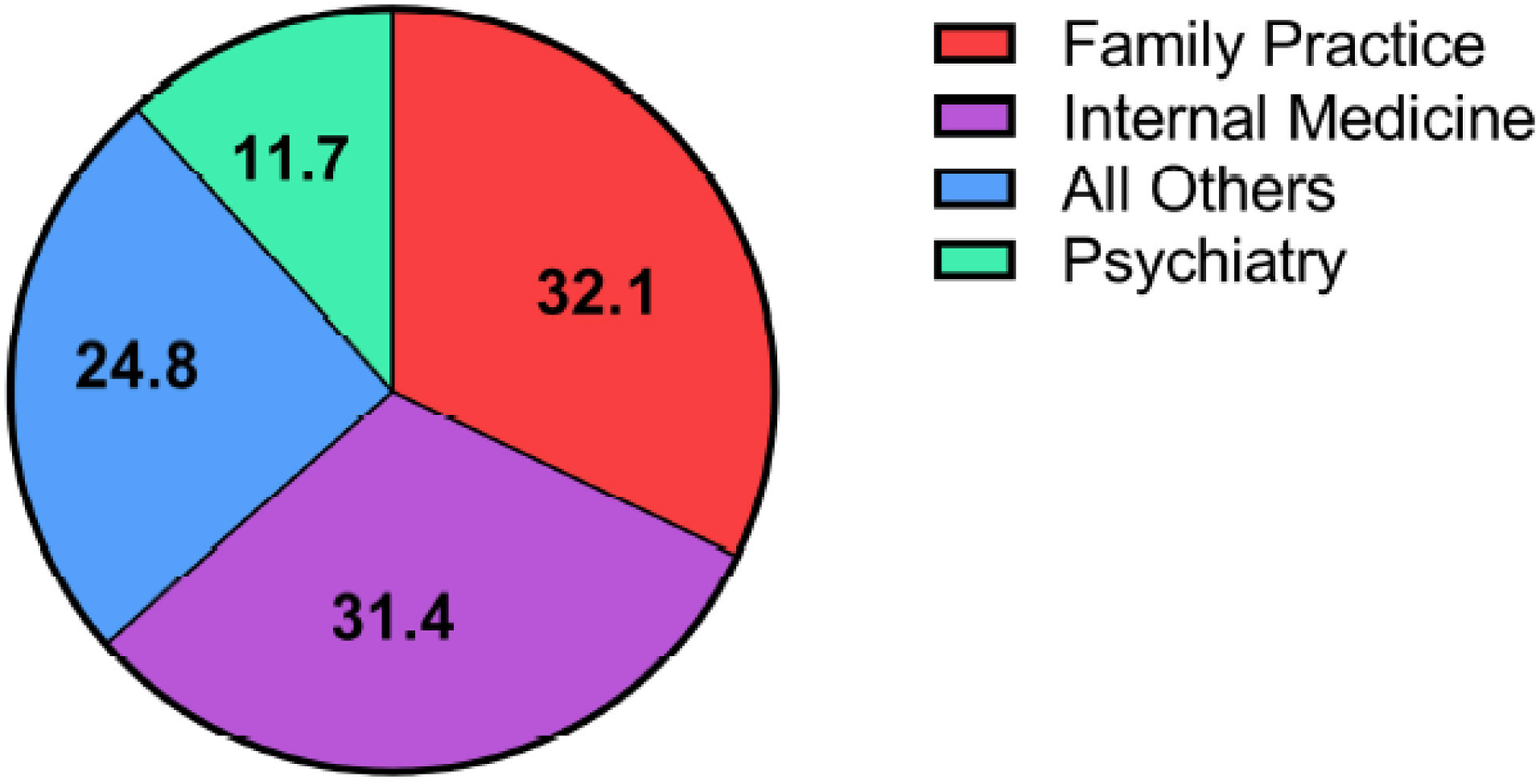
Z-drug days supply per prescription to Medicare enrollees by state.

**Supplemental Table 1.**
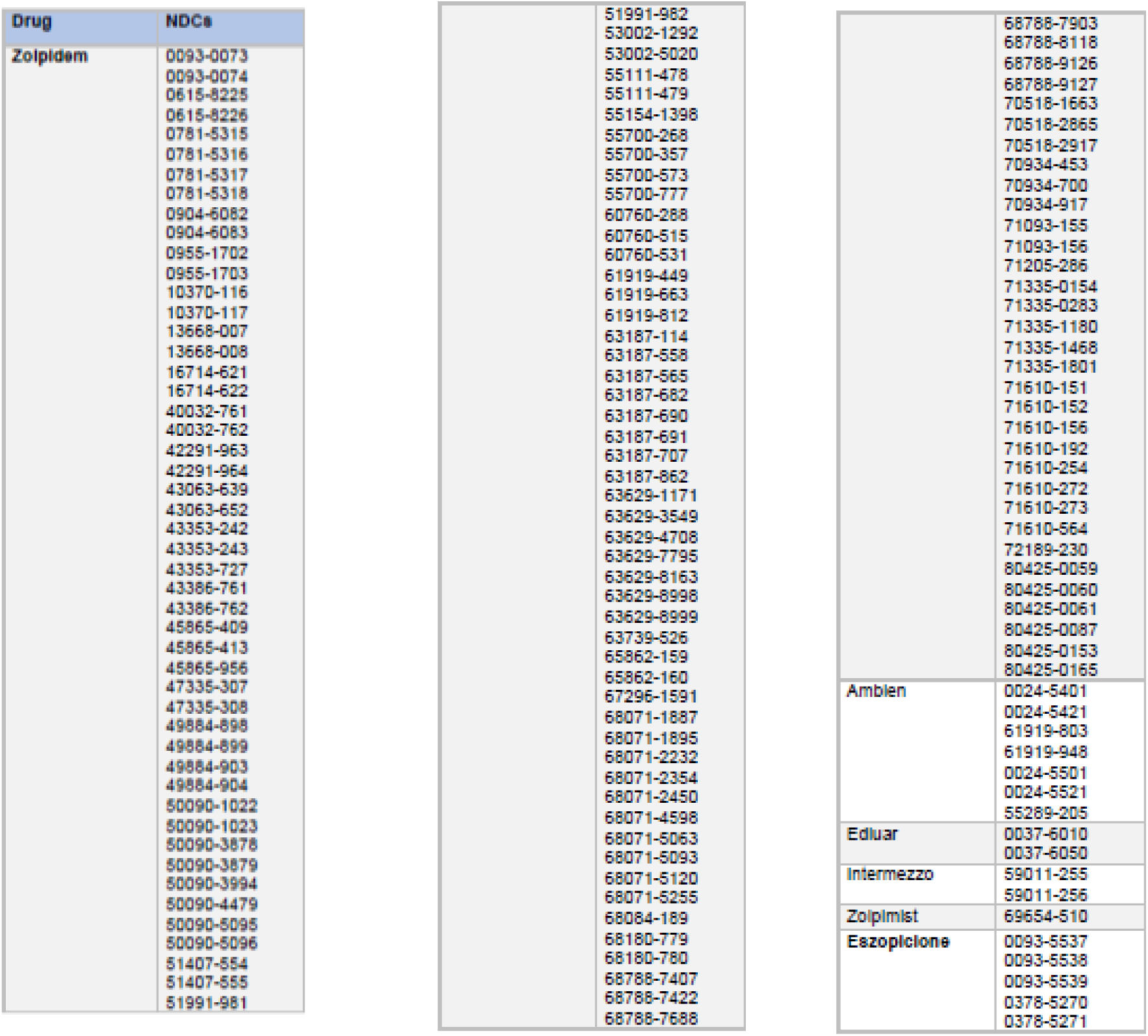
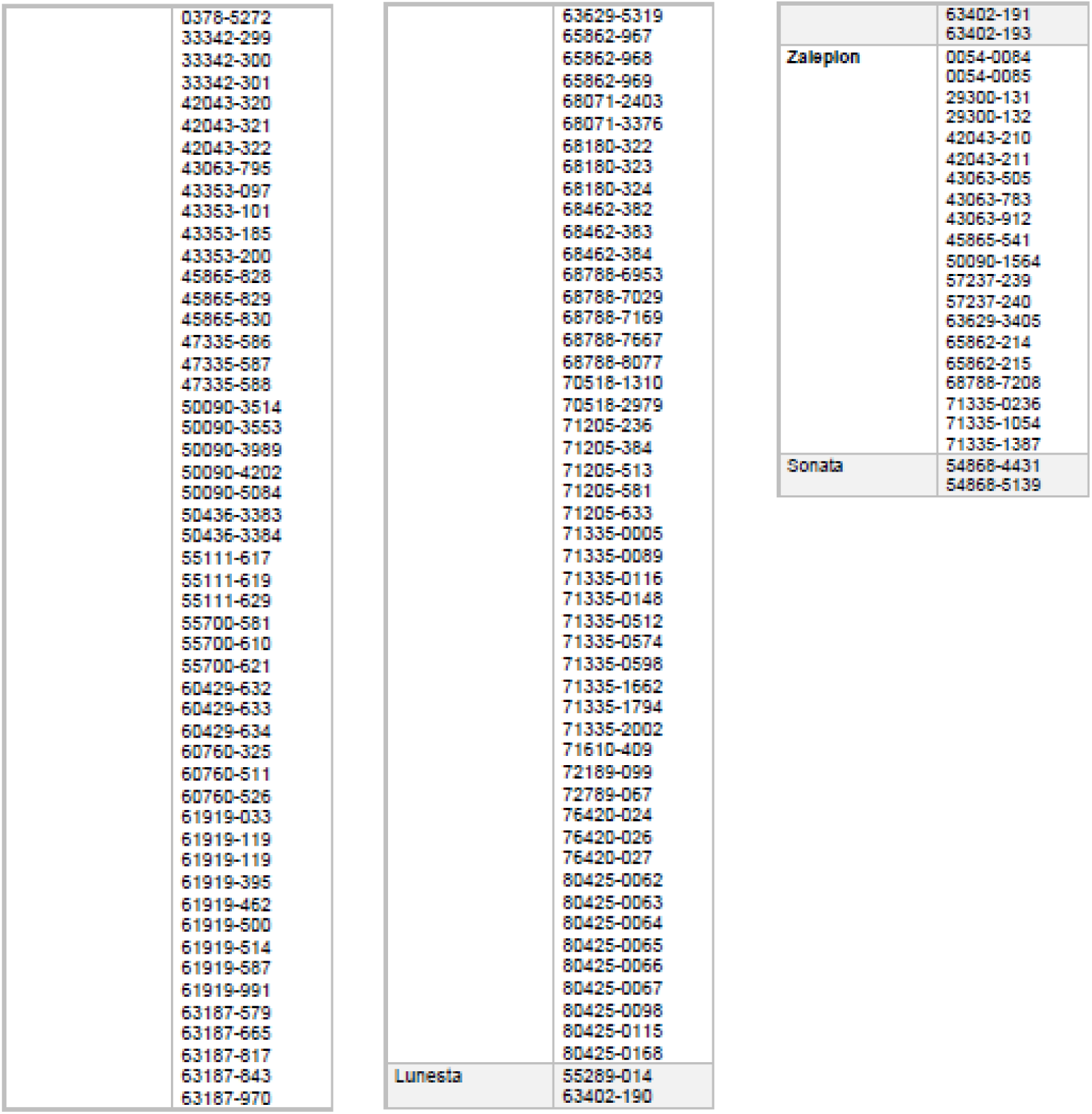
Z-drug generic (bold) and brand names with corresponding National Drug Codes (NDCs)

**Supplemental Appendix 1**. Python computer code to extract Medicare data

\# The purpose of this script is to pull info from Medicare provider .csv #

\# This needs to be run in the same directory as the .csv and as an “output.csv”

import csv

import pprint as pp

#format for the csv for providers

\# npi,nppes_provider_last_org_name, nppes_provider_first_name,

nppes_provider_city, nppes_provider_state, specialty_description,

description_flag, drug_name, generic_name, bene_count,

total_claim_count, total_30_day_fill_count, total_day_supply, total_drug_cost,

bene_count_ge65, bene_count_ge65_suppress_flag, total_claim_count_ge65,

ge65_suppress_flag, total_30_day_fill_count_ge65, total_day_supply_ge65,

total_drug_cost_ge65

file_name =

“Medicare_Provider_Utilization_and_Payment_Data 2018_Part_D_Prescriber.c

sv”

table_width = 0

ln_count = 0 all_data = {}

state_ls = [‘AE’,

‘AL’,

‘AK’,

‘AP’,

‘AZ’,

‘AR’,

‘CA’,

‘CO’,

‘CT’,

‘DC’,

‘DE’,

‘FL’,

‘GA’,

‘GU’,

‘HI’,

‘ID’,

‘IL’,

‘IN’,

‘IA’,

‘KS’,

‘KY’,

‘LA’,

‘ME’,

‘MD’,

‘MA’,

‘MI’,

‘MN’,

‘MS’,

‘MO’,

‘MT’,

‘NE’,

‘NV’,

‘NH’,

‘NJ’,

‘NM’,

‘NY’,

‘NC’,

‘ND’,

‘OH’,

‘OK’,

‘OR’,

‘PA’,

‘PR’,

‘RI’,

‘SC’,

‘SD’,

‘TN’,

‘TX’,

‘UT’,

‘VT’,

‘VA’,

‘VI’,

‘WA’,

‘WV’,

‘WI’,

‘WY’,

‘XX’,

‘ZZ’]

header = [“npi”,

“provider_lname”,

“provider_fname”,

“provider_city”,

“provider_state”,

“spec_desc”,

“desc_flag”,

“drug_name”,

“generic_name”,

“bene_count”,

“total_claim_count”,

“total_30_day_fill_count”,

“total_day_supply”,

“total_drug_cost”,

“bene_count_ge65”,

“bene_count_ge65_suppress_flag”,

“total_claim_count_ge65”,

“ge65_suppress_flag”,

“total_30_day_fill_count_ge65”,

“total_day_supply_ge65”,

“total_drug_cost_ge65”]

\# default encoding (UTF-8) throws an error - have to use ISO

\# Source has some nonstandard characters

with open(file_name, newline=‘‘, encoding=‘ISO_8859-1’) as csvfile:

dialect = csv.Sniffer().sniff(csvfile.read(1024))

csvfile.seek(0)

tempreader = csv.reader(csvfile, dialect)

#make the template row using the cols from above

template_output = {}

for col in header:

template_output[col] = ‘‘

for row in tempreader:

if ln_count == 0:

#This is just to process the header

table_width = len(row)

ln_count += 1

else:

i = 0 temp_row = {}

for col in header:

temp_row[col] = row[i]

i += 1

all_data[ln_count] = temp_row

ln_count += 1

\# all rows are in all_data now

\# essentially it’s the csv put into a python dict

#Get a list of unique providers so we can iterate through later

unique_provider = []

for state in state_ls:

for ln in all_data:

if all_data[ln][‘spec_desc’] not in

unique_provider: unique_provider.append(all_data[ln][‘spec_desc’])

else:

pass

#pp.pprint(unique_provider)

‘‘‘

\# Get totals for each specialty

num_providers = {} # {Family Practice: #, …}

for provider in unique_provider:

num_providers[provider] = 0

used_npi = []

for ln in all_data:

if all_data[ln][‘npi’] not in used_npi:

num_providers[all_data[ln][‘spec_desc’]] += 1

used_npi.append(all_data[ln][‘npi’])

else:

pass

pp.pprint(num_providers)

input()

‘‘‘

\# Broad scope looking at each provider perscribing as percent of total

spec_prescribing_output = {}

for spec in unique_provider:

spec_prescribing_output[spec] = 0

for ln in all_data:

temp_claim_count = all_data[ln][“total_claim_count”]

temp_claim_count = temp_claim_count.replace(‘,’,’’) #removed commas

ex. 1,800 -> 1800

spec_prescribing_output[all_data[ln][“spec_desc”]] +=

int(temp_claim_count) pp.pprint(spec_prescribing_output)

input()

progress_counter = 0

state_data = {} # ex. {PA:{spec1:claims,spec2:claims, … total:claims}}

for state in state_ls:

state_data[state] = {}

#build out list of provider types for each state

for provider in unique_provider:

state_data[state][provider] = 0

#tabulate total claims for each state

for ln in all_data:

temp_claim_count = 0

temp_claim_count = all_data[ln][“total_claim_count”]

temp_claim_count = temp_claim_count.replace(‘,’,’’) #removed commas

ex. 1,800 -> 1800

state_data[all_data[ln][“provider_state”]][all_data[ln][“spec_desc”]]

+= int(temp_claim_count) progress_counter += 1

if progress_counter % 1000 == 0:

print(progress_counter)

‘‘‘

#uncomment this if necessary - takes a while

#get number of providers per state

state_num_providers = {} # {‘PA’:100, ‘CT’: 50, …}

for state in state_data: state_num_providers[state] = 0

used_npi = []

for ln in all_data:

if all_data[ln][‘npi’] not in used_npi:

state_num_providers[all_data[ln][‘provider_state’]] += 1

used_npi.append(int(all_data[ln][‘npi’]))

else:

pass

pp.pprint(state_num_providers) ‘‘‘

‘‘‘

with open(‘output.csv’, mode=‘w’) as

csv_file: csv_writer = csv.writer(csv_file, delimiter=‘,’)

state_total = 0

for state in state_data.keys():

state_total = 0

for specialty in state_data[state]:

state_total += state_data[state][specialty]

csv_writer.writerow([state, specialty,

state_data[state][specialty]])

csv_writer.writerow([state, state + ‘ total’, state_total])

print(state_total)

print(“done”)

‘‘‘

‘‘‘

npi = row[0]

provider_lname = row[1]

provider_fname = row[2]

state = row[3]

spec = row[4]

desc_flag = row[5]

trade_name = row[6]

generic_name = row[7]

bene_count = row[8]

total_claim_count = row[9]

total_30_day_fill_count = row[10]

total_day_supply = row[11

total_drug_cost = row[12]

bene_count_ge65 = row[13]

bene_count_ge65_suppress_flag = row[14]

total_claim_count_ge65 = row[15]

ge65_suppress_flag = row[16]

total_30_day_fill_count_ge65 = row[17]

total_day_supply_ge65 = row[18]

total_drug_cost_ge65 = row[19]

‘‘‘

